# Hypertensive Disorders of Pregnancy and Primary Aldosteronism

**DOI:** 10.64898/2026.01.14.26343954

**Authors:** Stéfanie Parisien-La Salle, Arnaldo Ferrebus, Eva E Abel, Laura C Tsai, Andrew J Newman, Cheng Hsuan Tsai, Anand Vaidya, Jenifer M Brown

**Affiliations:** Center for Adrenal Disorders, Division of Endocrinology, Diabetes, and Hypertension, Brigham and Women’s Hospital and Harvard Medical School, Boston, MA; Division of Endocrinology, Department of Medicine, Centre Hospitalier de l’Université de Montréal, Université de Montréal, Montréal, QC, Canada; Division of Cardiovascular Medicine, Brigham and Women’s Hospital and Harvard Medical School, Boston, MA; Division of Cardiology, Department of Internal Medicine, National Taiwan University Hospital and National Taiwan University College of Medicine, Taipei, Taiwan; Primary Aldosteronism Center at National Taiwan University Hospital, Taipei, Taiwan

**Author notes:** <u>Corresponding Author</u>: Jenifer M. Brown, MD Brigham and Women’s Hospital Harvard Medical School 75 Francis St, Boston, MA, 02115. <u>Disclosures</u>: AV reports consulting fees unrelated to the contents of this work from Corcept Therapeutics, Mineralys, HRA Pharma, Moderna, SideraBio, Vertex, AstraZeneca. JMB reports consulting fees unrelated to the contents of this work from Recordati Rare Diseases and AstraZeneca. The other authors have no funding to disclose.

## Abstract

**Background:** Hypertensive disorders of pregnancy (HDP) affect up to 15% of pregnancies and are linked to adverse maternal and fetal outcomes. Primary aldosteronism (PA) affects up to 25% of hypertensive patients. We examined PA prevalence in women with prior HDP and its relationship to hypertension trajectory.

**Methods:** Adults from across the U.S.A. meeting guideline-recommended screening criteria for PA were prospectively tested. Women with a self-reported history of HDP completed a questionnaire examining the relationship between PA and hypertension trajectory.

**Results:** Of 330 hypertensive parous women (62.4 ± 9.8y; 32.1% non-white), 83 (25.2%) reported a history of HDP. Women with HDP were younger at hypertension diagnosis (38.8 vs. 47.9y; p <0.001). The prevalence of a positive PA test was similarly high in those with and without HDP (26.5% vs 32%; *p = 0.35*). Among women with HDP, 63 completed the follow-up questionnaire, of whom 15 (23.8%) tested positive for PA. Compared with PA-negative women, those with PA reported a higher proportion of pregnancies complicated by hypertension (76.5% vs. 60.9%, p = 0.11) and fetal complications (55.6% vs. 27.9%, p <0.01). Hypertension trajectories also differed: sustained hypertension, defined as persistently elevated blood pressure beyond the postpartum period, was nearly twice as frequent in women with a positive PA test (66.7 vs. 37.5%; p=0.047).

**Conclusion:** Over 25% of women with hypertension and a prior pregnancy screened positive for PA, highlighting its high prevalence, irrespective of history of HDP. Women with HDP remain at elevated cardiovascular risk, and PA may represent a targetable contributor.

## Introduction

Hypertensive disorders of pregnancy (HDP), which encompass gestational hypertension, chronic hypertension and preeclampsia/eclampsia, complicate up to 15% of pregnancies and are the second leading cause of maternal mortality (1–3). HDP also reflect healthcare disparities, with certain racial groups and individuals from lower income backgrounds experiencing a higher frequency of disease (2). The impact of HDP extends beyond pregnancy, with women with a history of HDP having an increased risk of future cardiometabolic disease, including sustained hypertension, myocardial infarction, stroke, heart failure, type 2 diabetes and chronic kidney disease (4). The pathophysiology of HDP is most likely multifactorial, involving both maternal and placental factors including pre-existing maternal comorbidities, genetic and immune factors, reproductive history, age and race (4).

With growing recognition of the high prevalence of primary aldosteronism (PA) as a cause of hypertension (5), recent guidelines now recommend testing all patients with hypertension for PA (6, 7). However, this condition is widely underdiagnosed in the general population (5, 8–10), including among women of reproductive age, in whom it may contribute to the pathophysiology of a subset of HDP (11). Identifying these cases of PA is critical, as the management of blood pressure and potassium differs in the context of PA, particularly after pregnancy, where aldosterone-targeted therapy can be introduced to reduce long-term cardiovascular risk (12). The aim of this study was to evaluate the frequency of positive testing for PA in women with a history of self-reported HDP and its association with hypertension trajectory by leveraging a nationwide PA screening cohort (13).

## Methods

Supporting data are available from the corresponding author upon reasonable request.

### Study Design

This study was a follow-up investigation building on a previously published, nationwide PA screening program (13). Adults who met one or more clinical guideline criteria for PA testing (14) were prospectively enrolled from locations across the United States between November 3, 2021, and July 1, 2024. Participants completed remote consent and a medical history review, including questions about pregnancy and HDP, followed by PA testing via blood draw at a local reference laboratory without interruption of medications. Women who endorsed a pregnancy history, with or without HDP, were included in this analysis as the primary study population. Those reporting a positive history of HDP were subsequently invited to complete a follow-up questionnaire to evaluate the relationship between a positive PA test result and pregnancy outcomes and hypertension trajectory (“HDP Cohort”).

### Study Population

As this study began in 2021, inclusion criteria reflected pre-2025 Endocrine Society clinical guideline recommendations (14, 15). Women were eligible for inclusion if they met one or more guideline criteria for PA testing, including: resistant hypertension, hypertension with spontaneous or diuretic-induced hypokalemia or potassium supplement prescription, hypertension with atrial fibrillation or atrial flutter, hypertension with adrenal mass, hypertension with obstructive sleep apnea, or hypertension with a family history of PA or of stroke or hypertension with onset prior to age 40y. Exclusion criteria were a previously established diagnosis of PA, a history of adrenalectomy, or current prescription of a mineralocorticoid receptor (MR) antagonist. All participants provided informed consent, and the study protocol was approved by the Mass General Brigham Institutional Review Board.

#### Self-reported history of HDP

Participants reported a history of prior pregnancy, and if present, reported any history of chronic hypertension, gestational hypertension, preeclampsia, or eclampsia during the medical history review.

#### PA screening status: positive vs. negative

Based on a previously published algorithm (13, 16, 17) designed to maximize sensitivity for possible PA, a positive PA test was defined as a PRA ≤ 1.0 ng/mL/h and an aldosterone concentration ≥ 5 ng/dL (both measured by LC-MS/MS), or PRA> 1.0 ng/mL/h and Aldosterone-to-Renin Ratio (ARR) ≥ 20. Conversely, a negative PA test was defined by aldosterone <5 ng/dL, or by PRA> 1.0 ng/mL/h with Aldosterone-to-Renin Ratio <20. A sensitivity analysis defined a positive PA test instead based on the newly released 2025 Endocrine Society PA guideline diagnostic cutoff of aldosterone ≥7.5 ng/dL by LC-MS/MS (6), rather than 5 ng/dL, in the context of a PRA ≤ 1.0 ng/mL/h.

#### HDP Cohort

Participants who endorsed a history of HDP on initial medical history review subsequently received an optional follow-up questionnaire (**Supplement material**) which ascertained the number and timing of all pregnancies. For each pregnancy, participants self-reported whether the pregnancy was impacted by HDP, the specific hypertensive disorder (i.e. gestational hypertension, preeclampsia, or eclampsia), as well as fetal complications, including pregnancy loss after 12 weeks, prematurity, and small-for-gestational-age births.

#### Hypertension Trajectories

Within the HDP cohort, we first evaluated hypertension outcomes using a two-trajectory framework based on the last hypertensive pregnancy:

- Resolved hypertension: blood pressure returned to normotensive levels by the end of the postpartum period, defined as 3 months after delivery.
- Sustained hypertension: persistent blood pressure elevation after the postpartum period.

Following this two-trajectory analysis, we further refined our characterization of blood pressure patterns by categorizing four trajectories based on the timing and persistence of elevated blood pressure across pregnancies and beyond the postpartum period. This inclusive approach allowed us to distinguish meaningful patterns, including when more than one pregnancy was impacted by HDP, which are known to represent different risk profiles (18, 19). Four trajectories were defined using self-reported blood pressure status during and after each pregnancy:

- Single Episode & Resolved: Hypertension in only one pregnancy, with return to normotensive blood pressure postpartum and throughout any subsequent pregnancies.
- Single Episode & Sustained: Hypertension in only one pregnancy, but persistently elevated blood pressure beyond the postpartum period.
- Recurrent & Resolved: Hypertension in more than one pregnancy, but normotensive blood pressure between pregnancies and after the postpartum period.
- Recurrent & Sustained: Hypertension present in multiple pregnancies, with persistently elevated blood pressure between pregnancies and/or after the postpartum period.

### Laboratory Assays

All laboratory assays were performed through the Quest Diagnostics commercial laboratory system, as previously described in detail (16, 20, 21). Plasma renin activity (PRA) was determined using LC-MS/MS to determine the rate of generation of angiotensin I, with a lower limit of quantitation of 0.10 ng/mL/h. Plasma aldosterone concentration was measured by LC-MS/MS, with a lower limit of quantitation of 1 ng/dL or 5 ng/dL, with values reported as <1 and <5 set equal to 0.99 and 4.99 ng/dL, respectively, for analytical purposes.

### Statistical Methods

In the overall study population, women with and without a history of HDP were compared to assess rates of PA and differences in clinical characteristics. Within the HDP cohort, further comparisons of pregnancy outcomes and hypertension trajectories were made between women with a positive versus negative PA test result. Baseline characteristics were reported as frequencies and percentages for categorical variables and as mean ± standard deviation (SD) or median with interquartile range (25^th^-75^th^ percentile) for continuous variables, depending on distribution. Continuous variables were compared using t-test for normally distributed data or the Wilcoxon rank-sum test for non-normally distributed variables. Categorical variables were compared using either Fisher’s exact test (for small frequencies) or the Chi-square test. A p-value of <0.05 was considered statistically significant. All statistical analyses were conducted using R with RStudio (Version 2024.12.1+563).

## Results

### Overall Study Population

A total of 330 women who reported at least one prior pregnancy were tested for PA as part of the nationwide, direct-to-patient testing program (13). Of these women, 83 (25.2%) reported a history HDP, while 247 (74.8%) did not. The overall rate of a positive test for PA was high at 30.6%, and rates did not differ significantly between women with and without HDP (26.5% vs. 32.0%, *p=0.35*) (**Figure 1**). The frequency of a positive PA test was slightly lower when the 2025 Endocrine Society guideline thresholds (6) were applied retrospectively (18.5% [61/330]) but remained similar regardless of HDP history: 21.7% (18/83) in women with a history of HDP versus 17.4% (43/247) in those without HDP (*p = 0.48*).

**Figure 1.**
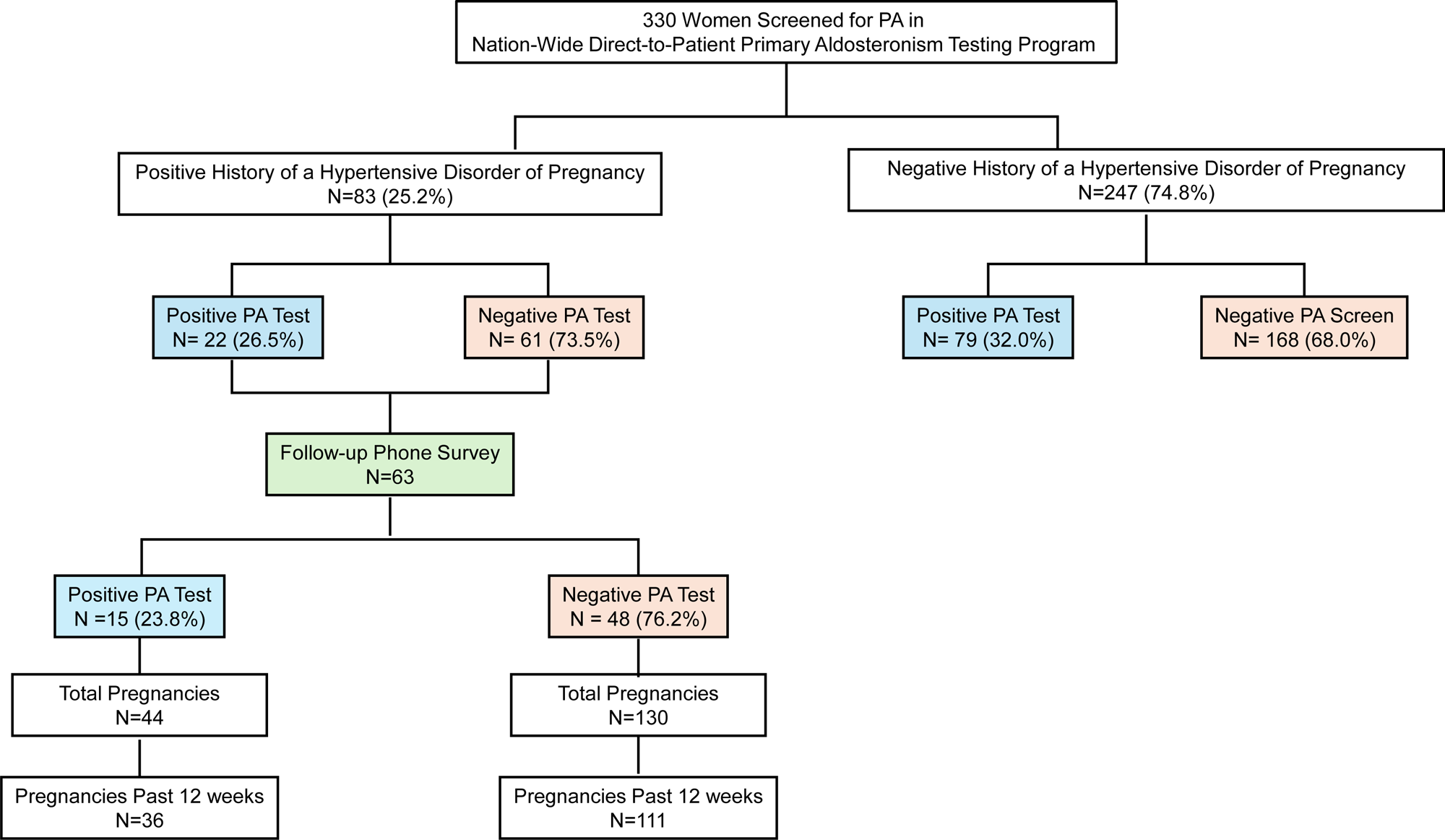
Study Flow Diagram. Flow diagram of women tested for primary aldosteronism (PA) in a nationwide, direct-to-patient PA testing program who reported at least one pregnancy, stratified by history of hypertensive disorder of pregnancy (HDP) and PA testing result.

Unsurprisingly, women with a history of HDP had an earlier age at hypertension diagnosis (38.8 ± 12.2 vs. 47.9 ± 11.2 years, *p* <0.001) and were significantly younger at the time of PA testing (mean age 57.6 ± 9.8 vs. 64.0 ± 9.2 years, *p* <0.001) than those without HDP. There were no significant differences in race, diabetes, chronic kidney disease, coronary artery disease, or sleep apnea between the groups. However, women with HDP were more likely to report continued usage of pregnancy-specific antihypertensives such as nifedipine or labetalol (12.0% vs. 3.2%, *p* = 0.002) almost 20 years following HDP, rather than more typical first-line antihypertensives (**Table 1**).

**Table 1.** Demographic and Clinical Characteristics of the Overall Study Population.

|  | Overall<br>n=330 | Categorical History of HDP |  | *p*-value |
| --- | --- | --- | --- | --- |
|  |  | Positive History of HDP<br>N=83 (25.2%) | Negative History of HDP<br>N=247 (74.8%) |  |
| **Age at PA testing, years** | 62.4 (9.8) | 57.6 (9.8) | 64.0 (9.2) | **< 0.001** |
| **Race, n (%)** |  |  |  |  |
| White | 224 (67.9) | 53 (63.9) | 171 (69.2) | 0.78 |
| Black | 50 (15.2) | 13 (15.7) | 37 (15.0) |  |
| Asian | 7 (2.1) | 2 (2.4) | 5 (2.0) |  |
| Other | 49 (14.8) | 15 (18.1) | 34 (13.8) |  |
| **Age at Hypertension Diagnosis, years** | 45.6 (12.1) | 38.8 (12.2) | 47.9 (11.2) | **< 0.001** |
| **Past Medical History, n (%)** |  |  |  |  |
| Diabetes | 63 (19.1) | 21 (25.3) | 42 (17.0) | 0.10 |
| Chronic kidney disease | 45 (13.7) | 14 (16.9) | 31 (12.6) | 0.33 |
| Coronary artery disease | 21 (6.4) | 5 (6.0) | 16 (6.5) | 0.92 |
| Sleep Apnea | 120 (36.4) | 29 (34.9) | 91 (36.8) | 0.76 |
| **Blood Pressure Medications, n (%)** |  |  |  |  |
| Calcium channel blockers | 138 (42.1) | 32 (38.6) | 106 (43.3) | 0.45 |
| ACE/ARBs | 226 (68.9) | 59 (71.1) | 167 (68.2) | 0.62 |
| Diuretics | 162 (49.4) | 43 (51.8) | 119 (48.6) | 0.61 |
| Beta-blockers | 107 (32.6) | 29 (34.9) | 78 (31.8) | 0.60 |
| **Pregnancy Specific Blood Pressure Medications, n (%)** |  |  |  |  |
| Nifedipine/Labetalol | 18 (5.5) | 10 (12.0) | 8 (3.2) | **0.002** |
| **GFR, mL/min/1.73m<sup>2</sup>** | 81.41 (20.63) | 86.1 (20.06) | 79.85 (20.62) | **0.016** |
| **Serum potassium, mmol/L** | 4.17 (0.4) | 4.14 (0.42) | 4.18 (0.39) | 0.44 |
| **Plasma Renin Activity, ng/mL/hr** | 1.58 (0.59-4.72) | 2.46 (0.72-7.43) | 1.49 (0.54-3.62) | **0.036** |
| **Plasma Aldosterone concentration, ng/dL** | 7 (4-11) | 8 (4.5-12) | 6 (4-10) | 0.068 |

Values are mean (SD), median (IQR), or n (%) for the overall population and stratified by self-reported history of hypertensive disorders of pregnancy (HDP). PA, primary aldosteronism; ACE, angiotensin-converting enzyme inhibitors; ARB, angiotensin receptor blocker; GFR, glomerular filtration rate.

### HDP Cohort

The optional follow-up phone survey was completed by 63 (75.9%) of the women with HDP. The frequency of PA in this cohort who provided HDP-related follow-up was consistent with the overall study population: 15 (23.8%) with a positive PA test and 48 (76.2%) with a negative test. If the updated 2025 PA guideline thresholds (6) were applied to the HDP cohort, only 3 participants would have been reclassified to PA negative, resulting in a positive PA test in 12 of 63 (19.0%).

These respondents reported 174 pregnancies in total, of which 147 progressed beyond 12 weeks gestation (36 in the PA-positive group and 111 in the PA-negative group) (**Figure 1**). Regarding population characteristics of the HDP Cohort, there were no significant differences in age at hypertension diagnosis or age at first hypertensive pregnancy between those testing positive or negative for PA (**Table 2**). Chronic kidney disease was significantly more common in women with a positive PA test (33.3% vs. 8.3%, *p* = 0.029). While not statistically significant, diabetes was numerically more frequent in the women with a positive vs. negative PA test (40.0% vs. 18.8%, *p=0.16*). First trimester pregnancy losses were also similar between both groups. The majority of pregnancies in the cohort were singleton, and among the small proportion of twin pregnancies (2.3%), there was no obvious association with PA status (**Table 2**).

**Table 2.** HDP Cohort: Patient and Pregnancy Characteristics.

| **<u>HDP Cohort</u>**<br>**<u>Patient Characteristics</u>** | **Categorical Classification of PA** |  |  |  |
| --- | --- | --- | --- | --- |
|  | **Overall**<br>**n=63** | **Positive PA Test**<br>**N = 15** | **Negative PA Test**<br>**N=48** | ***p-value*** |
| **Age at PA testing, years** | 58.68 (9.6) | 61 (8.0) | 57.96 (10.0) | 0.24 |
| **Age at hypertension diagnosis, years** | 38.9 (12.4) | 38.5 (14.86) | 39.0 (11.7) | 0.9 |
| **Age at first hypertensive pregnancy, years** | 27.2 (6.8) | 25.7 (9.6) | 27.6 (5.6) | 0.46 |
| **Past Medical History, n (%)** |  |  |  |  |
| Diabetes | 15 (23.8) | 6 (40.0) | 9 (18.8) | 0.16 |
| Chronic kidney disease | 9 (14.3) | 5 (33.3) | 4 (8.3) | 0.029 |
| Coronary artery disease | 4 (6.5) | 0 (0.0) | 4 (8.3) | 0.56 |
| Sleep Apnea | 22 (34.9) | 3 (20.0) | 19 (39.6) | 0.22 |
| **GFR, mL/min/1.73m<sup>2</sup>** | 86.8 (19.51) | 88.1 (19.12) | 86.4 (19.82) | 0.77 |
| **Serum potassium, mmol/L** | 4.13 (0.43) | 4.10 (0.35) | 4.14 (0.45) | 0.75 |
| **Plasma Renin Activity, ng/mL/hr** | 2.5 (0.82-8.02) | 0.59 (0.46-0.82) | 5.03 (1.8-12.17) | **<0.001** |
| **Plasma Aldosterone concentration, ng/dL** | 8 (4-12) | 10 (8-11.5) | 6 (3.75-12.25) | 0.08 |
| **Aldosterone to Renin ratio, ng/dL per ng/mL/hr** | 2.3 (1.3–8.9) | 16.4 (11.3-24.5) | 1.9 (1-3.1) | **<0.001** |
| **<u>HDP Cohort</u>**<br>**<u>Pregnancy Characteristics</u>** | **Categorical Classification of PA** |  |  |  |
|  | **Overall**<br>**n=174** | **Positive PA Test**<br>**N = 44** | **Negative PA Test**<br>**N=130** | ***p-value*** |
| **Number of pregnancy losses before 12 weeks\*, n (%)** | 25 (14.4) | 6 (13.6) | 19 (14.6) | 1.00 |
| **Number of twin pregnancies, mean (SD)** | 4 (2.3) | 2 (4.5) | 2 (1.5) | 0.25 |

Values are mean (SD), median (IQR), or n (%) for the full HDP cohort and stratified by PA test result. The upper panel describes the patient-level characteristics (n=63 individuals), and the lower panel describes pregnancy-level characteristics (n=174 pregnancies).
HDP, hypertensive disorders of pregnancy; PA, primary aldosteronism; GFR, glomerular filtration rate. \*2 additional patients experienced a pregnancy loss but did not recall exact timing, so were excluded from this analysis.

Given that blood pressure may not be consistently measured in all pregnancies prior to 12 weeks, rates of hypertension and complications were evaluated in only those pregnancies that progressed beyond 12 weeks gestation and are compared in **Figure 2**. Among these, a numerically greater proportion of hypertensive pregnancies were reported among women with a positive PA test compared to women with a negative PA test (76.5% vs. 60.9%; *p* = 0.106), while rates of preeclampsia were similar. In contrast, patient-reported fetal complications, including fetal losses after 12 weeks, prematurity, and small-for-gestational age, were significantly more frequent in women who tested positive for PA (55.6% vs. 27.9%, *p* <0.01).

**Figure 2.**
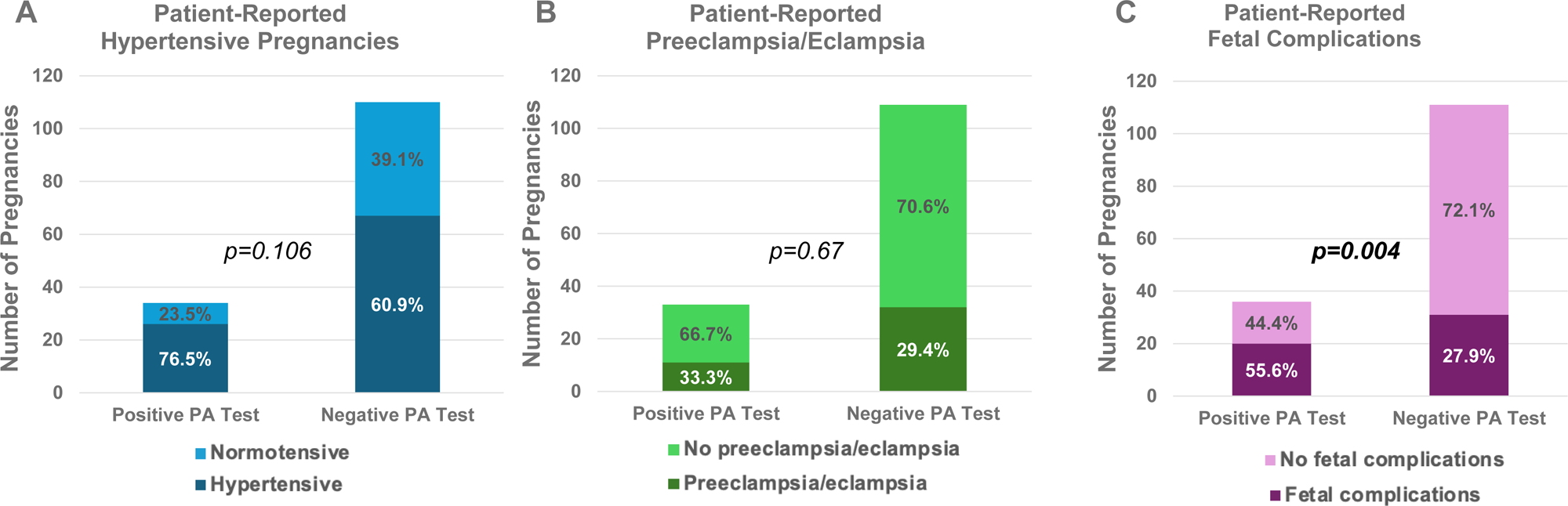
Pregnancy outcomes among women with positive versus negative PA testing results. Rates of self-reported obstetric and fetal complications: hypertensive pregnancies (A), preeclampsia/eclampsia (B) and fetal complications (C) in women with a history of HDP stratified by PA status. Fetal complications included pregnancy loss after 12 weeks, prematurity, and small-for-gestational-age births. Panel A: missing data for 3 pregnancies; Panel B: missing data for 5 pregnancies; Panel C: no missing data. HDP = hypertensive disorder of pregnancy; PA = primary aldosteronism.

Among women with a history of HDP, those who tested positive for PA exhibited less favorable hypertension trajectories. Sustained postpartum hypertension was twice as frequent in women with a positive PA test compared to a negative test (66.7% [10 of 15] vs 37.5% [18 of 48]; p = 0.047) (**Figure 3A and 3B**). We similarly observed a significant difference by PA status when examining the more detailed four-category trajectories (*p = 0.035*) (**Figure 3A and 3C**). Women with HDP who had a positive PA test were more likely to have an adverse hypertension trajectory, with higher rates of the “recurrent and sustained” pattern (60.0% [9/15] vs. 20.8% [10/48]). They were also less likely to follow a favorable trajectory, showing lower rates of the “single and resolved” pattern of blood pressure after HDP (33.3% [5/15] vs. 50.0% [24/48]) compared to women with a negative PA test.

**Figure 3.**
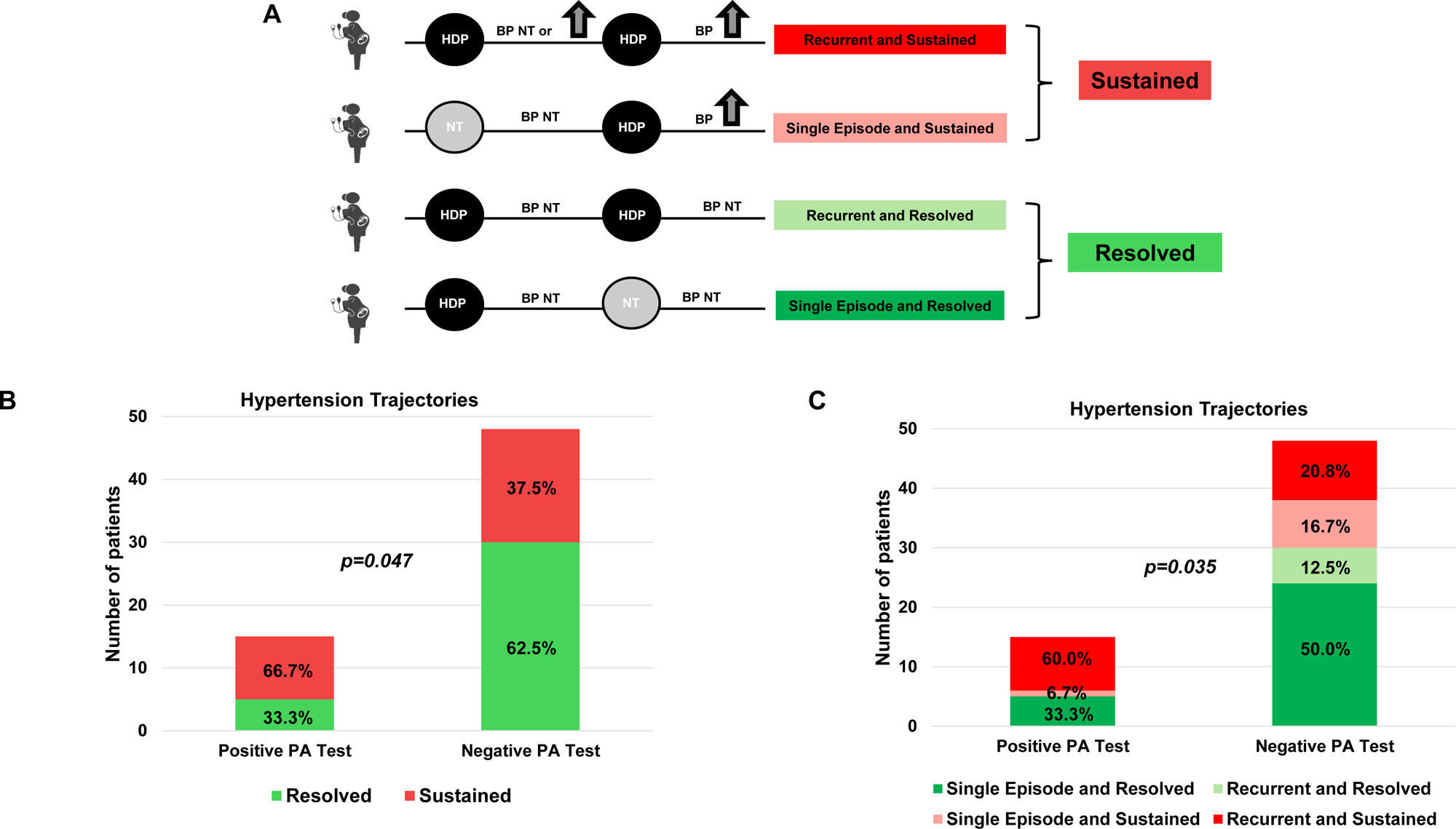
Hypertension trajectories among women with positive versus negative PA testing results. Hypertension trajectories were defined based on the number of episodes of HDP and whether blood pressure normalized postpartum (Panel A). The distribution of hypertension trajectories by PA status is shown in Panel B (sustained vs. resolved) and in Panel C (4 trajectories). BP = blood pressure; HDP = hypertensive disorder of pregnancy; NT = normotensive; PA = primary aldosteronism.

## Discussion

In this analysis of a nationwide cohort (13) that included 330 women with hypertension and at least one prior pregnancy, more than a quarter of women tested positive for suspected PA, with similarly high rates, though at younger ages, in those with a self-reported history of HDP. Among women with a history of HDP, a positive PA test was associated with more adverse self-reported pregnancy and hypertension outcomes, including more sustained hypertension, higher rates of fetal complications, and a trend toward a greater proportion of hypertensive pregnancies. These findings highlight that PA is highly prevalent among women with a history of pregnancy, irrespective of HDP, and may represent one mechanism contributing to more adverse cardiovascular outcomes in this at-risk population (11, 12).

A key finding of this study is the notably high overall prevalence of a positive PA test among hypertensive women (26.5% in women with a positive history of HDP and 32% in women without). When applying new guideline cutoffs (6) to our follow-up cohort (PAC ≥7.5 ng/dL in the context of suppressed PRA ≤ 1.0 ng/mL/h), the prevalence of PA remained high at 18.5%. Prior evidence on the rates of PA within HDP is of limited quality and predominantly based on observational reports reflecting clinical practice patterns without systematic PA testing (11). Reports of PA diagnosed in 3 of 2.8 million pregnancies (10) are likely marked underestimates, given consistently poor rates of testing in the general hypertensive population and the additional pregnancy-related challenges to making a PA diagnosis (8, 9, 22, 23), which likely contribute to the frequent lag between disease onset and diagnosis of PA, especially in women (24).

In our study, we also identified that women with a history of HDP and a positive PA test had a higher rate of self-reported fetal complications. This is consistent with small case series in which women with PA tended to have more complications than hypertensive non-PA patients (25, 26), and affirmed in a large systematic review by Sanga et al., where the overall complication rate, which included preterm delivery, fetal mortality, intrauterine growth restriction and preeclampsia was 62.2% in women with PA compared to 28.9% in women with essential hypertension (p <0.0001) (27), similar to what we report (55.6 vs 27.9% respectively). Whether recognition of PA before or during pregnancy could mitigate these adverse fetal outcomes is unknown. However, better pregnancy outcomes have been observed among individuals with normal preconception blood pressure (28, 29).

Within HDP, rates of preeclampsia were not associated with a positive test for PA in our cohort. The pathogenic link between PA and preeclampsia is not clearly established. On the one hand, women with preeclampsia have been shown to have a high prevalence of angiotensin II type 1 receptor autoantibodies (11, 30), which are also hypothesized to contribute to renin-independent aldosterone excess in PA (31), and preeclampsia has been associated with enhanced salt sensitivity and MR activation (32). Although preeclampsia rates were similar between PA groups, our study may have been underpowered to detect a difference given the limited number of affected pregnancies (n = 43). Conversely, preeclampsia is the result of a complex dynamic between placental-maternal-fetal factors (33), while PA results from a multi-hit process involving genetic susceptibility, paracrine signaling, aberrant receptors and altered receptor sensitivity (34–43). Thus, PA may instead play a greater role in prevalent chronic “essential” hypertension that manifests before or during pregnancy and is sustained postpartum, rather than in the pathogenesis of preeclampsia. For example, increased progesterone production during pregnancy can serve as an endogenous mineralocorticoid receptor antagonist, thus mitigating some of the blood pressure and vascular manifestations of PA during pregnancy that then may become more evident in the postpartum period after delivery of the placenta (25, 44). This phenomenon may be supported by our observation of more adverse blood pressure trajectories in the postpartum period among women with a positive PA test.

The clinical trajectory of PA across the reproductive lifespan has been reported with much variability. While in some cases, PA may be masked during pregnancy by the MR antagonist effect of progesterone (25, 44), in others, PA may worsen during pregnancy, leading to severe hypertension, adverse fetal outcomes, and, in rare instances, the need for adrenalectomy during the second trimester (25, 27). Thus, the clinical course of PA during pregnancy is difficult to predict. However, sustained blood pressure elevation after 12 weeks postpartum should prompt a work-up for secondary causes of hypertension (45). This was confirmed in a prospective study looking at overweight and obese participants with HDP, in which only 30% of participants with elevated blood pressure 2 months postpartum had normal blood pressure at 1 year after delivery (46). Also, future risk for chronic hypertension has been shown to be associated with the cumulative number of gestational hypertension/preeclampsia-affected pregnancies (19). Our findings support this association, as a positive PA test was linked to more hypertensive pregnancies and more sustained hypertension, suggesting a potential mechanistic basis for increased long-term risk. Indeed, chronic hypertension appears to account for approximately two thirds of HDP’s association with coronary artery disease and about half of its association with heart failure (47). The substantial burden of unrecognized PA in these already high-risk women is particularly important, given its impact on cardiovascular outcomes and modifiable nature (5, 48). Given the prevalence of PA approaching 25% or more of hypertension and the availability of targeted treatment, women with sustained hypertension postpartum or recurrent HDP warrant evaluation for PA. The persistence of nifedipine and labetalol use among women with HDP in our study demonstrates the pervasive clinical inertia impacting hypertension management in women with HDP and highlights a missed opportunity for disease-modifying treatment among those with PA.

### Strengths and Limitations

The main limitation of this study is its retrospective nature and reliance on self-reported data, which may introduce recall bias, and the relatively long interval between HDP and PA testing, which does not allow us to discern whether PA was present at the time of a hypertensive pregnancy or developed later in life. Also, the overall sample size was modest, with even smaller subgroups. Further, because the women tested for PA were in their 60s on average, obstetric care and management of HDP reflect older clinical practice. However, secular trends in standards of care have generally included more intensive monitoring and treatment of HDP(4, 49), and thus the more severe or overt cases of HDP were likely included in our cohort. Additionally, individuals with only one pregnancy or whose last pregnancy was the only one complicated by hypertension could not be classified as having “recurrent” hypertension and thus may have been misclassified as “resolved”. However, we included all participants to capture a broad range of real-life trajectories and identify clinically meaningful patterns, even among those with a single pregnancy. Regardless, misclassification of recurrent cases as resolved would have likely biased the results toward the nulI. Finally, participant inclusion was based on pre-2025 Endocrine Society guideline-recommended screening criteria for PA (14, 15). Consequently, some individuals who would have met the expanded screening recommendations of the 2025 guidelines (6) may not have been included in this study.

### Perspectives

More than 25% of women with hypertension and a history of pregnancy screened positive for PA, highlighting its high prevalence, irrespective of history of HDP. Women with HDP remain at increased risk for adverse cardiovascular outcomes, much of which is mediated through an increased burden of hypertension, and PA may represent one directly modifiable mechanism contributing to this trajectory. Given the long-term cardiovascular implications of HDP and the high prevalence of PA, this population should be considered for intensified PA testing in the postpartum period to determine candidacy for aldosterone-targeted interventions to mitigate cardiovascular risk.

### Novelty and Relevance

#### What Is New?

- More than 25% of hypertensive women with a prior pregnancy screened positive for primary aldosteronism, regardless of a history of hypertensive disorders of pregnancy.
- Among women with a history of hypertensive disorders of pregnancy, primary aldosteronism was associated with more self-reported fetal complications and more sustained hypertension postpartum.

#### What Is Relevant?

- Primary aldosteronism may represent a common and underrecognized contributor to hypertension in women with a history of hypertensive disorders of pregnancy.

#### Clinical/Pathophysiological Implications?

- Women with sustained hypertension after hypertensive disorders of pregnancy should be prioritized for primary aldosteronism screening.
- Targeted aldosterone-directed therapies may offer a strategy to reduce long-term cardiovascular risk following hypertensive disorders of pregnancy.

## Data Availability

Supporting data are available from the corresponding author upon reasonable request.

