## Supplemental Appendix for "Hypertensive Disorders of Pregnancy and Primary Aldosteronism"

### PA Pregnancy Questionnaire

Subject ID ______________________

How many times have you been pregnant?
- ◯ 0
- ◯ 1
- ◯ 2
- ◯ 3
- ◯ 4
- ◯ 5
- ◯ 6
- ◯ 7
- ◯ 8
- ◯ 9
- ◯ 10

#### Pregnancy 1

What was your age at the time of this pregnancy? ______________________

Did this pregnancy result in a live birth?
- ◯ Yes
- ◯ No

Did this loss occur after 12 weeks (3 months)?
- ◯ Yes
- ◯ No
- ◯ Unsure

Was this loss related to hypertensive complications?
- ◯ Yes
- ◯ No
- ◯ Unsure

How many babies resulted from this pregnancy?
- ◯ one
- ◯ two
- ◯ three
- ◯ four
- ◯ five
- ◯ six

Did you have hypertension during this pregnancy?
- ◯ Yes
- ◯ No
- ◯ Unsure

Which of the following characterized your hypertension during this pregnancy?
- ☐ High blood pressure
- ☐ Need for blood pressure medications
- ☐ Preeclampsia
- ☐ Other

Was hypertension diagnosed before pregnancy, during, or immediately after delivery?
- ◯ Before
- ◯ During
- ◯ Immediately after
- ◯ Unsure
- ◯ NA

During which trimester of this pregnancy was hypertension diagnosed?
- ◯ Trimester 1
- ◯ Trimester 2
- ◯ Trimester 3
- ◯ Unsure

| | |
|---|---|
| Did you need medications to manage BP before this pregnancy? | ◯ Yes<br>◯ No<br>◯ Unsure |
| How many medications did you need to manage your hypertension before this pregnancy? | ◯ 1<br>◯ 2<br>◯ 3<br>◯ 4<br>◯ 5<br>◯ 6 |
| Did you need medications to manage BP during this pregnancy? | ◯ Yes<br>◯ No<br>◯ Unsure |
| How many medications did you need to manage your hypertension during this pregnancy? | ◯ 1<br>◯ 2<br>◯ 3<br>◯ 4<br>◯ 5<br>◯ 6 |
| Did you need medications to manage BP after this pregnancy? | ◯ Yes<br>◯ No<br>◯ Unsure |
| How many medications did you need to manage your hypertension after this pregnancy? | ◯ 1<br>◯ 2<br>◯ 3<br>◯ 4<br>◯ 5<br>◯ 6 |
| Did you have low potassium levels during pregnancy? | ◯ Yes<br>◯ No<br>◯ Unsure |
| Did you take a potassium supplement during this pregnancy? | ◯ Yes<br>◯ No<br>◯ Unsure |
| Were you diagnosed with "preeclampsia" or "eclampsia" during this pregnancy? | ◯ Yes<br>◯ No<br>◯ Unsure |
| Were you diagnosed with "preeclampsia" or "eclampsia" during this pregnancy? | ◯ Preeclampsia<br>◯ Eclampsia |
| Did you have any kidney, liver, blood count, seizures, or other organ complications that you know of? | ______________________________ |
| Were you delivered early during this pregnancy? | ◯ Yes<br>◯ No<br>◯ Unsure |
| Was this early delivery because of high blood pressure? | ◯ Yes<br>◯ No<br>◯ Unsure |

| | |
|---|---|
| Did you have any other complications during this pregnancy? | ◯ Yes<br>◯ No<br>◯ Unsure |
| What were these other complications? | __________________________ |
| Did the baby have any complications such as prematurity or growth restriction/small for gestational age? | ☐ No<br>☐ Prematurity<br>☐ Growth restriction/small for gestational age<br>☐ Other |
| Other: | __________________________ |
| Did your blood pressure go back to normal after this pregnancy? | ◯ Yes<br>◯ No<br>◯ Unsure |
| Comments: | __________________________ |

**Other**

| | |
|---|---|
| At what age were you ultimately diagnosed with hypertension? | __________________________ |
